# A Framework to Monitor Editing of Artificial Intelligence-Generated Medical Documentation

**DOI:** 10.64898/2026.09.30.26364427

**Authors:** Augusto Garcia-Agundez, Siyu Zhou, Jessica Pourian, Catherine Blebea, Parnaz Daneshpajouhnejad, Elizabeth Dente, Kevin Shi, Sarah Pollet, Fan Xia, Xu Shi, Robert Thombley, Cynthia Fenton, Sara G. Murray, Julia Adler-Milstein, Gabriela Schmajuk, Jean Feng, Jinoos Yazdany

**Affiliations:** Department of Medicine, University of California, San Francisco, CA USA; University of Michigan, Ann Arbor, MI USA; San Francisco VA Medical Center, San Francisco, CA USA

**Keywords:** AI, NLP, Clinical Documentation, AI Monitoring, AI Safety

## Abstract

**Objective:** To develop and evaluate a framework for characterizing clinician editing of Artificial Intelligence (AI)-generated documentation and assess its feasibility for health system-level monitoring of AI scribes.

**Materials and Methods:** We analyzed outpatient encounters in which an AI scribe was used at a single academic health system, examining the History of Present Illness (HPI) and Assessment and Plan (A&P) sections of notes. We characterized edits on three dimensions: lexical edit intensity based on Levenshtein distance, embedding edit intensity BERTScore, and clinical edit intensity based on removed and added UMLS concepts. We analyzed the Positive Predictive Value (PPV) of clinical edit intensity as a measure of clinically meaningful editing using clinicians as the gold standard, examined correlations among dimensions, and designed exponentially weighted moving-average control charts to monitor longitudinal changes in clinician editing behavior.

**Results:** 268,379 encounters were included (267,654 HPI, 267,594 A&P). Clinical edit intensity ≥1 had an 88.9% PPV for clinically meaningful editing. Lexical and embedding edit intensity were highly correlated (Spearman ρ 0.95), while clinical edit intensity was less strongly correlated with both (ρ 0.77-0.81). Longitudinal monitoring detected changes coinciding with system-wide rollout.

**Discussion:** Clinicians edited A&Ps more heavily than HPIs, potentially reflecting greater attention to content involving clinical decision-making. Over one-third of sections involved clinical concept changes, and clinical edit intensity identified clinically meaningful edits while providing information complementary to lexical editing measures.

**Conclusion:** Clinician editing can be characterized at scale using complementary editing dimensions, providing a scalable signal for post-deployment surveillance of the human-AI documentation process.

## BACKGROUND AND SIGNIFICANCE

Clinical documentation drives physician burnout, reduces time available for direct patient care, and causes workflow inefficiency across health systems.[1] Generative artificial intelligence (GenAI) has emerged as a potential solution, assisting with tasks such as drafting clinical documentation.[2] Adoption has been rapid, and hundreds of health systems now use AI scribes.[2–11] Early evaluations report reductions in documentation time[12, 13] and burnout,[6] improvements in clinician satisfaction[5] and billing,[14] and favorable effects on patient experience.[15]

AI scribes differ from other AI decision-support tools: they generate free-text for the medical record, with the clinician as the sole checkpoint between model output and a legally and clinically consequential document. The rapid deployment of clinical GenAI has outpaced the development of monitoring methods. Unlike prior AI tools, for which post-deployment surveillance relies on well-established accuracy metrics,[16] GenAI scribes lack an established human-in-the-loop oversight framework. Available measures of editing focus on the likelihood or amount of AI-generated text in documents,[17] not what changed, or how meaning may have changed as a result.

AI scribe literature has emphasized adoption,[2–5, 7, 9–14] satisfaction,[2–10] time savings,[2–5, 7, 8, 10–13] and revenue impact,[3, 10, 14] with little attention to how clinicians interact with model outputs. Recent work has highlighted clinician modification of AI-generated drafts as a critical yet understudied dimension, indicating the need for methods to systematically characterize these interactions.[18] Preliminary evidence indicates that AI scribes can introduce errors during transcription and drafting,[19] and that cognitive surrender[20] and automation bias,[21] well-documented phenomena in other human-AI interactions, are risks in routine clinical use. Qualitative studies reveal some reasons for editing: revise factual discrepancies, add details or diagnostic reasoning, convert patient expressions into objective assessments, and reorganize or rephrase content.[22]

Clinician edits of AI-generated drafts signal whether the AI system is functioning as intended and whether meaningful oversight is being exercised. However, how to monitor and interpret these edits remains unclear. Minimal editing may reflect a good AI draft or clinician overreliance. Extensive editing may reflect careful oversight or a poor draft. Exactly what was edited may also provide insights: whether the clinician corrected a small section, rephrased existing content, added new content, or modified a mention of a diagnosis or procedure. Scalable, computationally efficient measures of clinician editing could provide useful signals for post-deployment monitoring, particularly if they capture clinically meaningful changes.

## OBJECTIVE

In this study, we assembled a dataset of over 250,000 clinical encounters with AI-scribed content across over 1,000 clinicians over more than 20 months of deployment. We evaluated candidate measures of lexical, embedding, and clinical edit intensity to determine: (1) their validity and complementary information, and (2) their utility as scalable surveillance signals for post-deployment monitoring of AI performance and clinician oversight.

## MATERIALS AND METHODS

### Data source, study period, and scribe workflow

Data were derived from outpatient clinical encounters July 2024-March 2026 at a large academic center using Epic as its Electronic Health Record (EHR) and Abridge^1^ as its AI scribe. This study was approved by the center’s IRB.^2^

For each encounter, we collected: (1) AI-Generated Sections, which were the initial AI-generated note sections, for the History of Present Illness (HPI) and Assessment and Plan (A&P) sections of the sampled clinical notes; (2) Clinician-Edited Sections, which were the section text after any edits by human authors, if available; and (3) the full text of the complete, signed clinical note. We also collected encounter characteristics used in stratified analyses, including visit type (new patient vs. follow-up) and interpreter requirement (yes vs. no/unknown). We studied the HPI and A&P because they were the most-requested sections and contain the encounter’s clinical narrative and reasoning.

### Inclusion/Exclusion Criteria

We included all ambulatory encounters during the study period with at least one AI-Generated Section.

We excluded (1) encounters in which a scribe other than Abridge was used, and (2) encounters without an AI-Generated Section for the analyzed section (HPI or A&P).

### Definitions of Scribe Use

A Clinician-Edited Section is created only when an AI-Generated Section is imported via a supported workflow (e.g., dotphrase or template) and edited. However, its absence does not necessarily indicate that the AI-generated Section was unused: A clinician may also import it through an alternative workflow (e.g., copy-paste), use it without editing, or not use it. We therefore developed methods to infer how AI-generated content was used for each section and encounter.

We used string searches in the final note for the Clinician-Edited Section and AI-Generated Section, after removing non-alphanumeric characters to account for EHR-induced formatting changes, to determine whether their content was incorporated. Possible clinician workflows and preprocessing steps in our framework are depicted in **Figure 1**.

**Figure 1:**
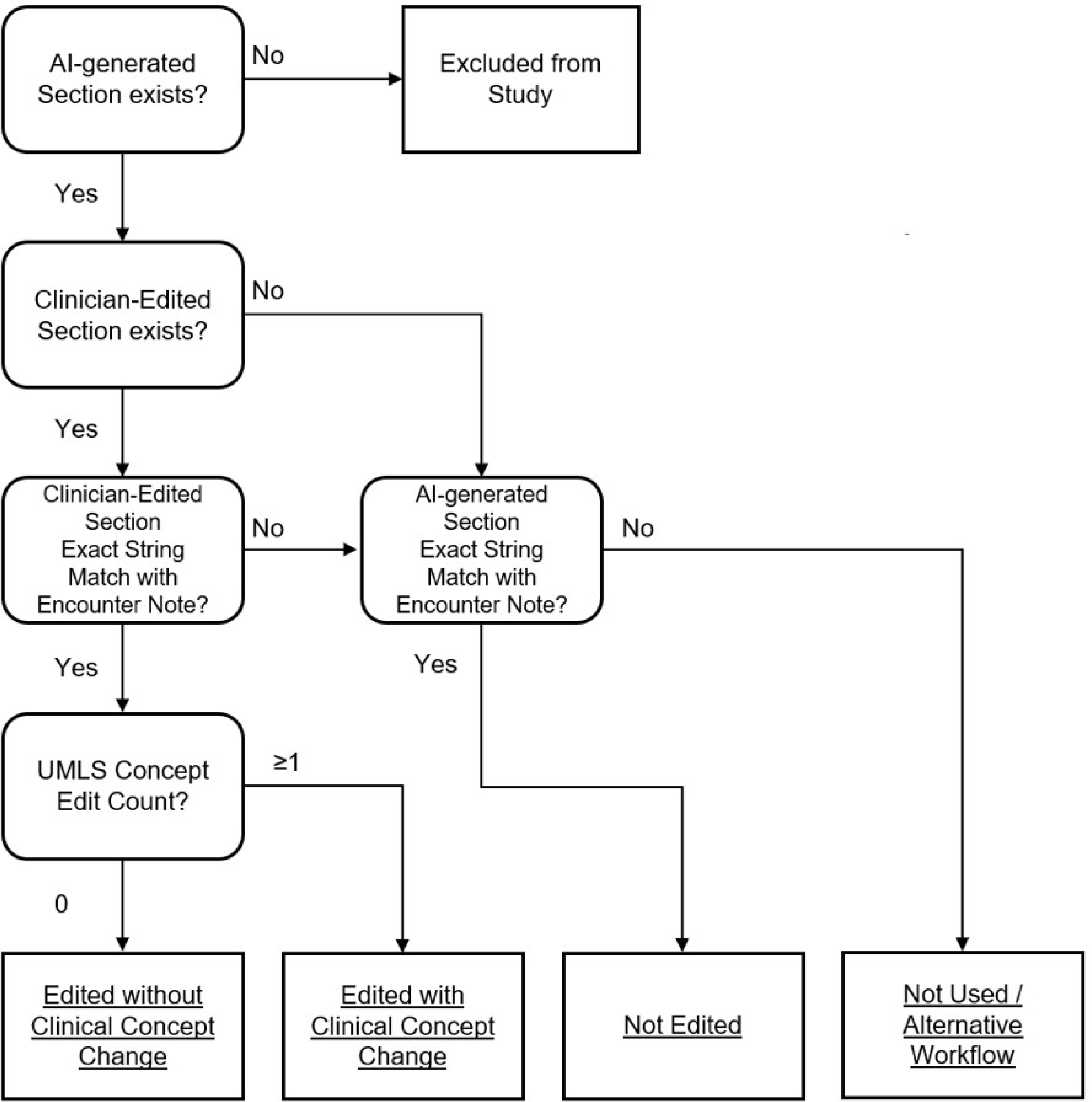
Study diagram of clinician workflows and resulting editing profile categories. This diagram is applied separately to each note section, i.e., the History of Present Illness (HPI) and Assessment and Plan (A&P). Each section was assigned to an editing profile based on whether the edits included clinical concept changes and whether the AI-Generated Section was used verbatim Alt text: Flow diagram showing the classification of AI-generated clinical note sections into four editing profiles. Sections without AI-generated content are excluded. Sections with clinician-edited content are assessed for clinical concept changes using UMLS concept edits and classified as edited with or without clinical concept change. Sections without clinician-edited content are classified as not edited when the AI-generated text exactly matches the final note, or as not used/alternative workflow when no exact match is found.

### Framework Edit Dimensions

We developed a framework for evaluating AI-generated clinical documentation using three candidate dimensions of editing, each operationalized with an established metric from the Natural Language Processing (NLP) or clinical literature. The framework integrates existing measures to characterize complementary aspects of editing AI-generated text. We prioritized measures that could be computed automatically and efficiently at scale. We selected lexical and embedding edit intensity because these represent two commonly studied dimensions in NLP, and clinical edit intensity because prior work proposed incorporating a clinical dimension into analyses of editing.[23] These dimensions may capture overlapping aspects of clinician editing, and we evaluated the extent of this overlap empirically. **Table 1** shows examples of edits for each dimension based on edits observed in our data.

**Table 1.**
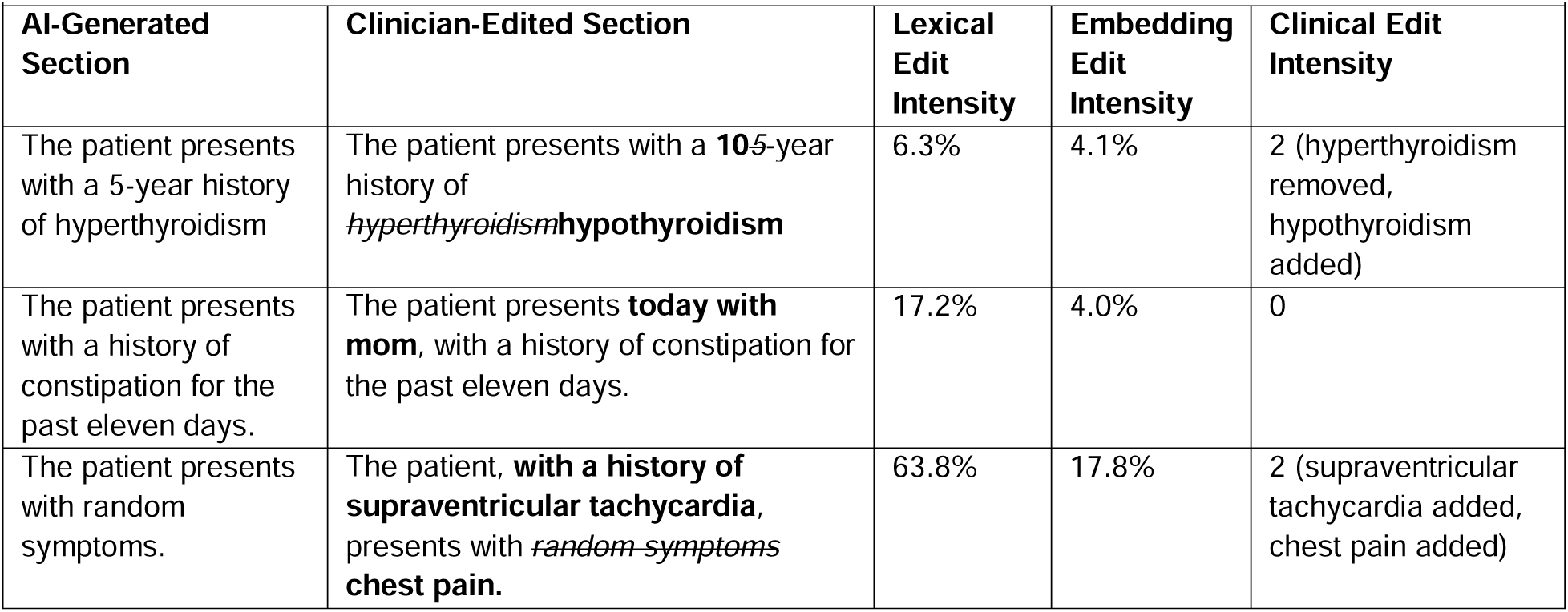
Examples of lexical, embedding, and clinical edit intensity values for different Sections. The AI-Generated Section is the text originally proposed by the AI scribe, while the Clinician-Edited Section is the same text after the clinician’s edits are complete. Text proposed by the AI and removed by the clinician is depicted in. New text proposed by the clinician is depicted in **bold**.

Our first dimension, **lexical edit intensity**, measured character-level edits, i.e., how much text has changed, regardless of changes in meaning. We selected the Normalized Levenshtein Edit Distance using Yujian and Bo’s formula,[24] a well-defined measure of surface-level editing that yields a continuous [0,1] variable, with higher values indicating more editing. Other candidates considered, both from NLP literature and proposed by us, included BLEU, ROUGE-L, the Jaccard token index (using TikToken’s GPT-4 vocabulary), and Edit Density, defined as the number of individual edits divided by the section length. We elected to use Levenshtein Edit Distance because related work supports its use to measure clinician edits of AI-generated text.[23]

Our second dimension, **embedding edit intensity**, used embedding-based methods to assess how text has changed. Because embeddings encode semantic information, we hypothesized that changes in embedding space would help capture shifts in meaning. We selected section-level BERTScore,[25] which has been extensively validated, correlates better with human judgment than similar measures such as cosine similarity, and has previously been proposed for assessing clinical textual similarity.[23] We used a clinical encoder (BioClinical-ModernBERT-large) expecting that training on clinical data would make it more sensitive to changes in clinical meaning. We normalized the measure to follow the same continuous order as lexical edit intensity by calculating 1 , so that a higher value means more editing.

Our third dimension, **clinical edit intensity**, detected when edits add or remove clinical concepts. It was proposed in prior work[23] because neither lexical nor embedding edit intensity capture changes in clinical meaning: embedding-based methods, in particular, can assign a low embedding difference to clinically opposed yet semantically related concepts, thereby understating a substantial change in clinical meaning. Prior work has attempted to mitigate this by combining an embedding similarity metric with a clinical ontology database, but has not demonstrated improved capture of changes in clinical meaning.[23] Clinical edit intensity counts removed or added Unified Medical Language System (UMLS) concepts, comparing the set of UMLS concepts identified in the AI-Generated Section and the Clinician-Edited Section. This approach was selected because UMLS concepts are widely used in clinical NLP.[26] Concepts present only in the AI-Generated Section were defined as removed; concepts present only in the Clinician-Edited Section were defined as added. Changes in the frequency of concepts were ignored. Clinical edit intensity is represented as the sum of removed and added concepts, each contributing one point. This metric was not normalized because each concept may represent a discrete inaccuracy, omission, or hallucination, regardless of visit complexity.

### Framework Edit Profiles

Based on clinical edit intensity, we defined four mutually exclusive editing profiles (**Figure 1**), defined as: 1. Edited without Clinical Concept Change (CCC); 2. Edited with CCC; 3. Not Edited; and 4. Not used/Alternative Workflow.

Sections that were edited and had zero clinical edit intensity (i.e., no UMLS concepts were removed or added) were defined as edited without CCC. This profile may include stylistic edits, rephrasing, reorganization, grammar correction, or other text-level changes. Sections with clinical edit intensity ≥1 (i.e., ≥1 UMLS concept removed or added) were defined as edited with CCC. This profile may include corrections of clinical inaccuracies, additions of omitted content, removal of irrelevant or unsupported content, or other edits involving clinical concepts.

If the AI-Generated Section was found verbatim in the final note, we categorized that profile as Not Edited. If our string-matching algorithm found neither the AI-Generated nor the Clinician-Edited Section in the final note, we categorized it as Not Used/Alternative Workflow, i.e. encounters in which either an alternative workflow (e.g., copy-paste) was used or the AI-Generated Section was not used at all. We report the proportion of sections matched by each search and treat the not used/alternative workflow profile as a residual category.

### Analyses

Our analyses aimed to evaluate the validity, interpretability, and feasibility of our framework for characterizing and monitoring clinicians’ editing of AI-generated drafts. Applying the framework to the data described above, we conducted three complementary analyses:

#### Validation of clinical edit intensity as a measure of clinically meaningful edits

A key component of our framework is that clinical edit intensity can distinguish clinically meaningful edits. To evaluate the criterion validity of our threshold of clinical edit intensity ≥1, we compared it against a clinician-adjudicated reference classification. We selected 54 HPIs and 54 A&Ps, with equal numbers having clinical edit intensity 0 and ≥1, as the smallest number allowing balanced allocation across sampling strata. Sections were sampled to represent variation in editing dimensions and encounter characteristics. Four clinicians blinded to clinical edit intensity independently reviewed 27 HPIs and 27 A&Ps each, where two clinicians reviewed each section, and each annotator pair overlapped in 9 HPIs and 9 A&Ps. Annotators judged whether the edits in each section were clinically meaningful. Disagreements were adjudicated by a panel of two senior clinicians to establish the reference classification. Treating this adjudication as the reference standard, and including 95% confidence intervals (CI) for all outcomes, our primary outcome was the positive predictive value (PPV) of clinical edit intensity ≥1 to detect clinically meaningful editing. Our secondary outcomes were the negative predictive value (NPV) of clinical edit intensity 0, the overall percent agreement and Cohen’s κ between clinical edit intensity classification and the reference standard, and median and range of inter-rater percent agreement and κ. Annotator instructions and inter-annotator agreement prior to adjudication for all annotator pairs, as well as PPV for various thresholds of clinical edit intensity , and a breakdown of results for HPIs and A&Ps, are reported in the Supplementary Material.

#### Analysis of the relationship among editing dimensions

Whether lexical, embedding, and clinical edit intensity capture similar or distinct aspects of clinician editing is unknown. To assess their association, we calculated pairwise Spearman correlations among the three edit dimensions separately for HPIs and A&Ps. We additionally calculated correlations between normalized Levenshtein edit distance and other candidates for lexical edit intensity described before.

#### Evaluation of the framework for monitoring system-level changes in editing

A central aim of our framework was to enable monitoring of health system-level editing trends for process changes that may warrant investigation. To assess the feasibility of this application, we implemented exponentially weighted moving average (EWMA) control charts. EWMA control charts have been proposed for process change detection in clinical AI applications.[16] To evaluate whether the EWMA could identify a known system-level change in editing behavior, we used the transition from pilot testing to scribe rollout to all outpatient providers as an *a priori* determined control event.

EWMA run charts followed the methodology described by Qiu.[27] For each edit dimension, we calculated the weekly mean across encounters with edits. For each editing profile, we calculated the weekly proportion of all eligible encounters in each profile. The first eight weeks of data were considered a warm-up period. We constructed weekly EWMA control charts using 0.75, 2.3, corresponding to 50. We selected this set of variables as a balance between robustness and sensitivity. We report the total number of alarms, and which edit dimensions and profiles exceeded the EWMA control limits due to the control event.

To assess the performance of the EWMA outside of the known control event, and to characterize the sensitivity of the proposed values of λ and ρ, we generated prospective simulations through the end of the study period to quantify the magnitude of change required to exceed the EWMA control limits. Our primary outcome was the magnitude of the minimum relative change needed to exceed the control limit.

Additionally, to examine behavior across strata of interest and overall EWMA performance, control charts for new patient encounters and encounters requiring an interpreter, control charts using other values of λ and ρ, and prospective EWMA control limits for different values of λ and ρ, were calculated and are provided in the Supplementary Material. All statistical analyses were performed in Python.

## RESULTS

The study included 268,379 encounters from 142,235 unique patients and 1,081 providers. AI-generated HPI and A&P sections were available for 99.7% of encounters. **Table 2** summarizes encounter characteristics, editing dimensions, and editing profiles. Overall, HPIs underwent less extensive lexical, embedding, and clinical editing than A&Ps. CCCs were common in both sections, occurring in more than one-third of encounters. HPIs were more frequently incorporated without editing than A&Ps (37.3% vs 19.3%) and less frequently classified as not used or incorporated through an alternative workflow (14.6% vs 33.8%). These patterns were generally similar across encounter characteristics.

**Table 2:**
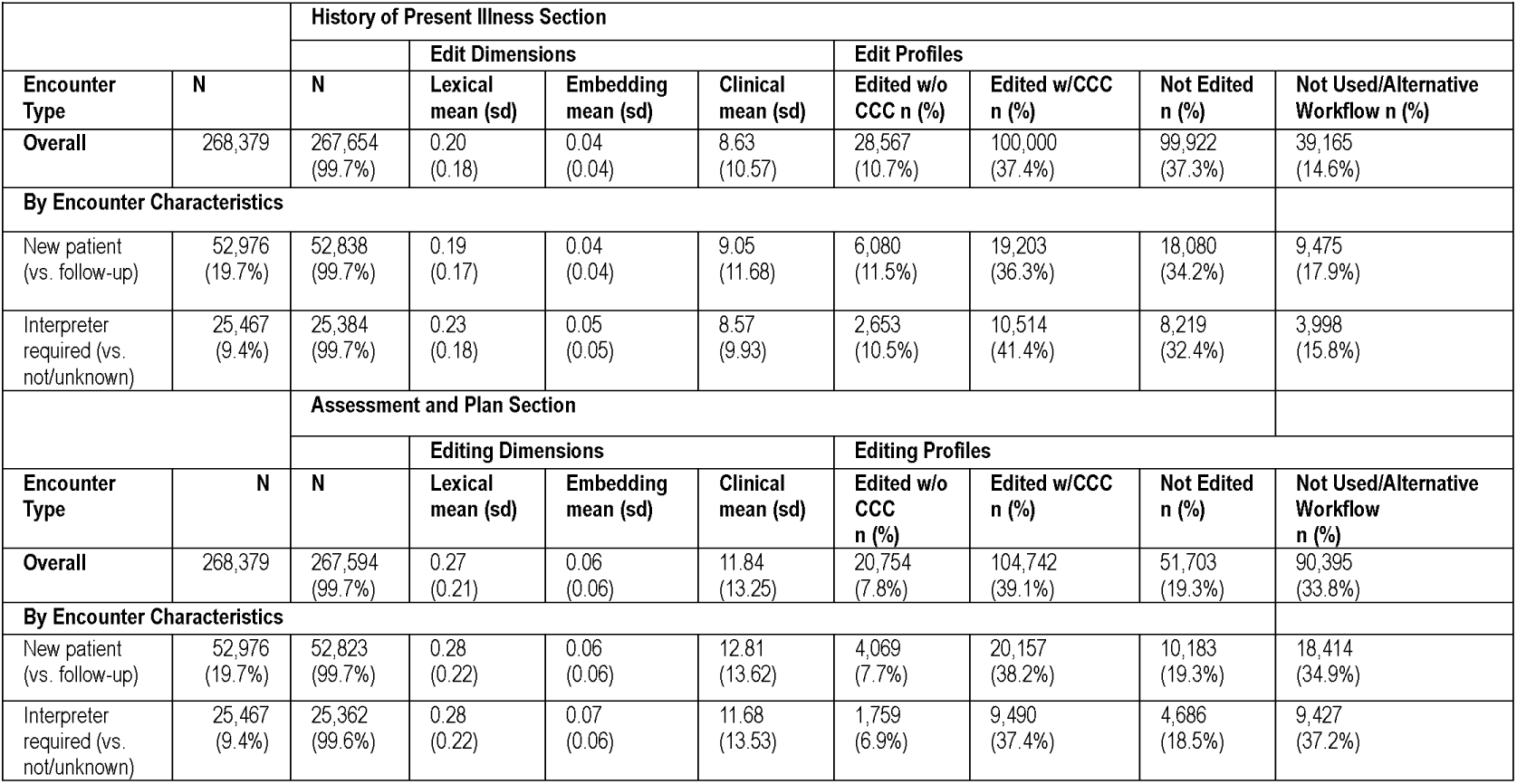
Encounter characteristics. CCC: Clinical Concept Change, sd: Standard Deviation. The N for the history of present illness and assessment and plan sections refers to the number of encounters that had an AI-Generated Section overall, and as the percentage of total encounters. Lexical, embedding, and clinical refer to lexical, embedding, and clinical edit intensity, respectively, and are calculated among encounters with sections classified as edited with or without CCC. Description of each editing profile presented in Figure 1.

### Validation of clinical edit intensity as a measure of clinically meaningful edits

The PPV of clinical edit intensity ≥1 to detect clinically meaningful edits was 88.9% (95% CI 77.8-94.8%). The NPV of clinical edit intensity 0 was 68.5% (95% CI 55.3-79.3%). Overall percent agreement between clinical edit intensity and the reference standard was 78.7% (95% CI 70.1-85.4%), with a Cohen’s κ of 0.57 (95% CI 0.42-0.73). Clinician annotators demonstrated moderate agreement in determining whether edits were clinically meaningful. The inter-rater median percent agreement was 66.7% (range 61.1%-88.9%). The inter-rater median κ was 0.33 (range 0.28-0.73). Inter-annotator agreement prior to adjudication for all annotator pairs, as well as PPV for different thresholds of clinical edit intensity other than ≥1, and a breakdown of results for HPIs and A&Ps, are reported in the Supplementary Material.

### Analysis of the relationship among editing dimensions

Lexical and embedding edit intensity were highly correlated in both HPIs A&Ps (Spearman ρ=0.95 for both), suggesting that they largely overlap. Clinical edit intensity was less strongly correlated with lexical (Spearman ρ=0.77 HPI, 0.80 A&P) and embedding edit intensity (Spearman ρ=0.78 and 0.81), supporting its use as a complementary measure of changes in clinical content. Alternative measures of lexical edit intensity were also highly correlated with our selected measure (Spearman ρ=0.93-0.98). Given the substantial overlap between lexical and embedding edit intensities, we retained lexical edit intensity for subsequent analyses because it was less computationally intensive and included clinical edit intensity as a complementary measure.

### Evaluation of the framework for monitoring system-level changes in editing

**Figure 2** shows weekly EWMA control charts for the two final edit-intensity dimensions and four editing profiles, separately for HPIs and A&Ps. We observed 16 alarms in total over 51 weeks and 12 signals. Three alarms were associated with the control event. For the HPI, the Not Edited profile was below control limits for two weeks, starting the week after rollout, while the Not Used profile was above control limits for one week, five weeks after rollout. For the A&P, the Not Edited profile was above control limits for one week, the week of the rollout.

**Figure 2:**
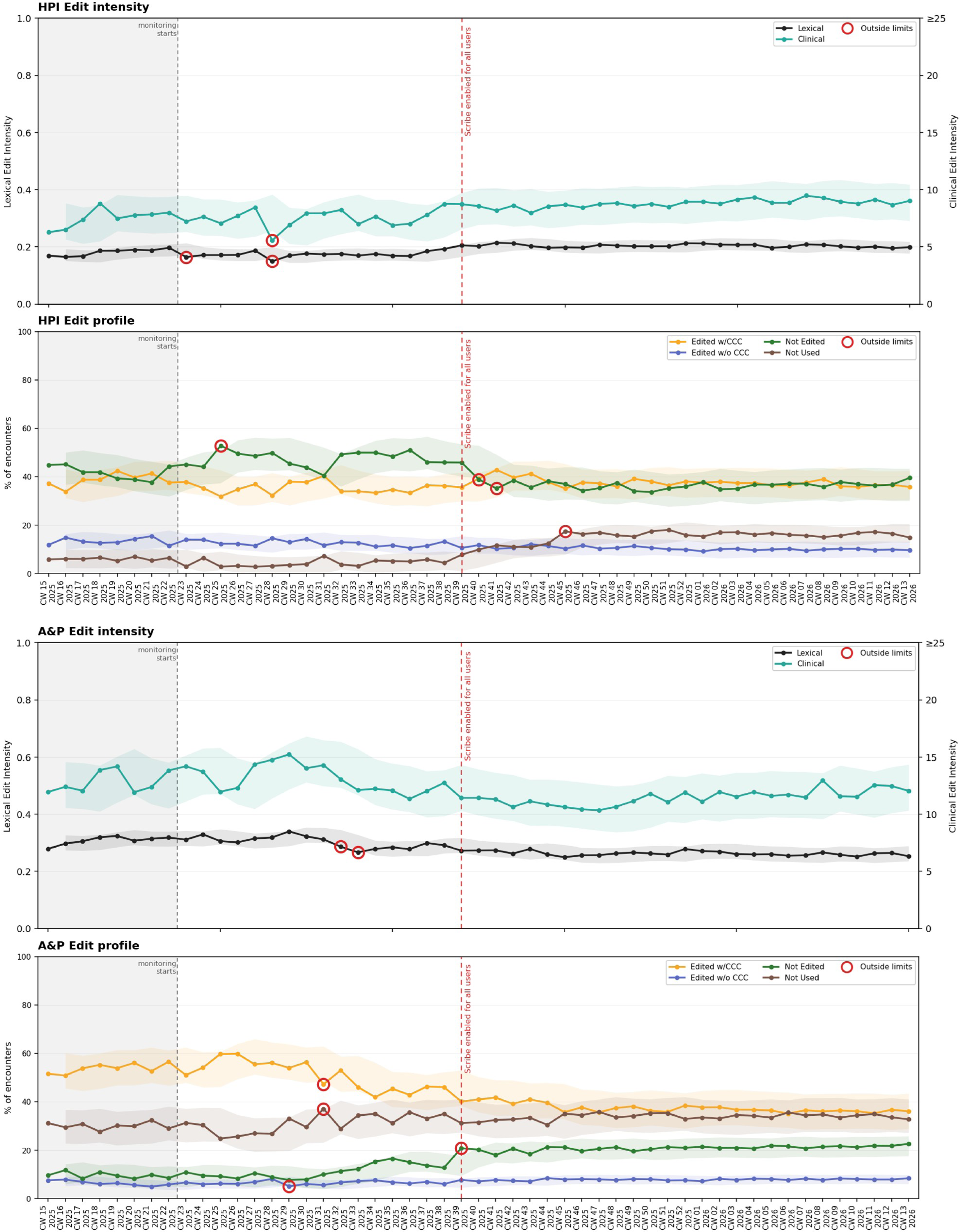
Weekly Exponentially Weighted Moving Average (EWMA) run charts of framework edit intensities and profiles at the health system level across history of present illness (HPI) and assessment and plan (A&P) sections at our academic health center. The unit of measure in the x axis is calendar week (CW). Values exceeding the EWMA thresholds are circled in red. The red, vertical dashed line indicates a known changepoint: the week in which the scribe was released to clinicians for all outpatient encounters. We consider the first eight weeks of data a warm-up period. ‘Not Used’ includes all encounters in the ‘Not used/Alternative Workflow’ profile. Run chart parameters: 0.75, 2.3. CCC: Clinical Concept Change. Alt text: Weekly exponentially weighted moving average control charts showing lexical and clinical edit intensity and four editing profiles for History of Present Illness and Assessment and Plan sections. Control limits are shown around each time series, with points exceeding the limits marked as alarms. A vertical dashed line indicates system-wide AI scribe rollout. Several alarms occur around rollout, including changes in the not-edited and not-used profiles, while most subsequent weekly values remain within control limits

Prospective simulations quantified the magnitude of the shift required to exceed the selected EWMA control limits. Changes of approximately 10% in Lexical or 15% in Clinical Edit Intensity were sufficient to exceed limits. For the profiles, the required change varied widely, from 14%-32% in the HPI to 16%-19% in the A&P.

In supplementary analyses, monitoring performance was generally less sensitive among encounter subgroups, and λ = 0.2 led to a very high alarm rate. Control charts for new patient encounters and encounters requiring an interpreter, charts using other values of λ and ρ, and prospective EWMA control limits for different values of λ and ρ are provided in the Supplementary Material.

## DISCUSSION

In this study of more than 268,000 clinical encounters involving an AI scribe, we found that clinician editing behavior can be characterized at scale and used as a signal for post-deployment monitoring of AI-generated clinical documentation. Our study has three important findings. First, clinicians frequently edited AI drafts, with heavier editing of the A&P than of the HPI, likely reflecting greater attention to content involving clinical decision-making. Second, over a third of sections involved CCCs, and clinical edit intensity ≥1 had an 89% PPV for detecting clinically meaningful edits and provided information complementary to lexical editing. Third, longitudinal monitoring of these measures in our health system detected a system-level change in scribe use associated with system-wide rollout. This work provides a framework for converting clinician interactions with AI-generated drafts into measurable signals that may support surveillance of generative AI after clinical deployment.

Clinician interaction with AI-generated documentation appeared to vary with the clinical stakes of the content. AI-generated HPIs, which primarily summarize the patient interview, were substantially more likely to be incorporated without editing. In contrast, A&Ps, which more directly reflect clinical interpretation and decision-making, were edited with CCCs more often. This likely suggests greater clinician surveillance as documentation moves from recounting the encounter toward clinical interpretation and reasoning. Alternatively, heavier A&P editing may reflect lower draft quality due to incomplete clinical context or limitations in clinical reasoning. Prior qualitative work similarly found that clinicians modify AI-generated drafts to correct factual discrepancies, add diagnostic reasoning, and transform patient-reported information into clinical assessments.[18] Our findings extend this by showing that clinician engagement with AI-generated documentation is not uniform across a note and may reflect the clinical consequences of the content. Post-deployment monitoring strategies that aggregate editing behavior across an entire note may therefore obscure differences in clinician interaction with higher- and lower-stakes content. Therefore, for health system monitoring, we have chosen to disaggregate note sections.

Our clinician annotation analysis supports using clinical edit intensity as a proxy for clinically meaningful editing, with high PPV for clinician-adjudicated annotations. However, NPV was lower: edits in nearly one-third of sections with clinical edit intensity 0 were judged clinically meaningful. Moderate inter-annotator agreement also suggests that determining whether an edit is clinically meaningful is itself a matter of clinical judgment. Annotator comments indicated that missed meaningful edits primarily involved modifying concept attributes and rearrangement. Clinical edit intensity should therefore be interpreted as a specific but partially sensitive measure of clinically meaningful editing. This limits its utility for individual note assessment, but may matter less at the health system level.

The lower correlation between Clinical and lexical edit intensity indicated that the extent of textual editing does not fully capture whether edits involve clinical concepts. Examples in our data illustrate this: replacing one diagnosis with another (‘hyperthyroidism’ to ‘hypothyroidism’) yields little lexical change despite altering clinical meaning, whereas adding contextual language (‘patient here today with mom’) produces greater lexical change without altering identified clinical concepts. This distinction is particularly relevant from a safety perspective, as small textual changes can substantially alter clinical meaning. Thus, despite its limitations, clinical edit intensity may capture clinically consequential editing that lexical measures alone can obscure. We also found that embedding-based measures did not provide additional information about editing beyond what lexical and clinical edit intensity can provide, and therefore we elected to retain only lexical and clinical edit intensity in our framework for monitoring purposes. Evaluating multiple candidate dimensions allowed us to identify a parsimonious set of measures that can be calculated automatically at scale for longitudinal monitoring. This may be informative for groups building similar pipelines since it argues against default use of embedding-based measures and enables cost-effective monitoring without LLMs.

Beyond characterizing individual edits, our longitudinal analysis demonstrated the feasibility of using clinician editing behavior for health system-level surveillance. EWMA monitoring detected a change in editing behavior coinciding with the transition from pilot testing to system-wide rollout. Following the rollout, newly eligible clinicians were more likely to edit the HPI and less likely to edit the A&P than clinicians who participated in the pilot. One plausible explanation is that pilot clinicians were tasked with actively evaluating the tool and likely to be self-selected AI adopters, whereas clinicians adopting the scribe after rollout used it in routine clinical practice. Detecting this expected shift provides evidence that editing-based measures can identify system-level changes in clinicians’ interactions with AI-generated documentation.

The control charts produced a few potentially false alarms, and none were felt to represent true process changes. We therefore characterized the magnitude of change required to trigger an alarm with the current EWMA values. Our analysis suggests that substantial relative changes are needed to exceed our proposed EWMA control limits. Alert thresholds should balance monitoring sensitivity with the health system’s AI governance capacity for review.

Signals identified outside the known rollout event illustrate the central challenge of routine surveillance: editing patterns can identify changes but cannot establish cause. Decreased editing may reflect improved AI-generated drafts that require fewer corrections or reduced clinician oversight, while increased editing may reflect deterioration in AI-generated drafts, shifting documentation preferences, or more intensive review. Editing behavior should be interpreted as a surveillance signal rather than a direct measure of AI performance or clinician oversight. The purpose of post-deployment monitoring is not to establish an optimal level of editing, but to detect meaningful departures from local patterns that warrant investigation. The sensitivity of editing surveillance should be calibrated to the types of changes a health system considers important and its capacity to investigate resulting signals.[28]

Our study has several limitations. First, clinician editing behavior changes can provide a high-level signal of potential model drift but cannot establish its cause. Changes in clinician behavior, workflows, patient populations, scribe model updates, or provider-level personalization could produce similar signals; detected changes, therefore, require further investigation. Second, clinical edit intensity captures additions and removals of identified clinical concepts but may miss clinically meaningful changes involving attributes such as numbers, doses, laterality, certainty, or temporality, as well as uncommon abbreviations or concepts with typographical errors. However, AI scribes are unlikely to commit typographical errors, making UMLS matching more reliable than expected. Third, comparisons between HPI and A&P editing may be subject to selection bias because clinicians used AI-generated A&Ps less frequently. Fourth, the Not Used/Alternative Workflow profile cannot distinguish abandonment of AI-generated content from use through an unsupported workflow, which may bias estimates of editing behavior if alternative workflows differ systematically in editing intensity. Fifth, our EWMA analysis is limited to one control event, and more work is needed to determine its sensitivity to detect changes of different magnitudes. Finally, this single-center study evaluated one AI scribe platform. Validation across health systems and AI scribes is needed to establish generalizability and determine which editing signals are most informative for detecting clinically important changes in model performance. Availability of the necessary editing data may also vary across platforms.

## CONCLUSION

We developed and evaluated a framework to characterize clinician editing of AI-generated clinical documentation across lexical and clinical dimensions and applied it to over 250,000 encounters at an academic health center. Our findings demonstrate the feasibility of using clinician editing behavior as a post-deployment monitoring signal, including characterizing clinically meaningful edits and detecting system-level changes in editing patterns. Future work should further validate and refine these measures across clinical settings and evaluate the actionability of editing signals and their impact on patient care.

## DATA AVAILABILITY STATEMENT

The data analyzed in this study contain protected health information (PHI) and therefore cannot be made publicly available due to privacy and confidentiality requirements. Python code to calculate editing intensities and profiles, and to generate EWMA charts used for the analyses is publicly available at https://github.com/AugustoGarcia/ai_scribe_editing_framework.

## AI USE STATEMENT

During the preparation of this work, the authors used Claude 5.5 in high-effort mode (Anthropic PBC, San Francisco, CA) for manuscript drafting, and GPT 5.5 in instant-effort mode (OpenAI Group PBC, San Francisco, CA) through the HIPAA-compliant platform at our academic center for code drafting. All AI-generated content was reviewed, edited, and verified by the authors for correctness. The authors take full responsibility for the final content of the manuscript and associated code.

## Footnotes

1 Abridge AI INC (San Francisco, USA)

2 The Institutional Review Board of the University of California, San Francisco approved this study and waived the requirement for informed consent (protocol number: 24-43295)

## REFERENCES

1 Moy AJ, Schwartz JM, Chen R, et al. Measurement of clinical documentation burden among physicians and nurses using electronic health records: a scoping review. J. Am. Med. Inform. Assoc. 2021;28:998–1008.

2 Olson KD, Meeker D, Troup M, et al. Use of ambient ai scribes to reduce administrative burden and professional burnout. *JAMA Netw*. Open 2025;8:e2534976.

3 Shah SJ, Garcia P. Ambient ai scribes—what is the return on investment? JAMA Netw. Open 2026;9:e2553238.

4 Tierney AA, Gayre G, Hoberman B, et al. Ambient artificial intelligence scribes: learnings after 1 year and over 2.5 million uses. NEJM Catal. 2025;6:CAT.25.0040.

5 Duggan MJ, Gervase J, Schoenbaum A, et al. Clinician experiences with ambient scribe technology to assist with documentation burden and efficiency. *JAMA Netw*. Open 2025;8:e2460637.

6 Albrecht M, Shanks D, Shah T, et al. Enhancing clinical documentation with ambient artificial intelligence: a quality improvement survey assessing clinician perspectives on work burden, burnout, and job satisfaction. JAMIA Open 2025;8:ooaf013.

7 Stults CD, Deng S, Martinez MC, et al. Evaluation of an ambient artificial intelligence documentation platform for clinicians. *JAMA Netw*. Open 2025;8:e258614.

8 Guo Y, Wang J, Hu D, et al. Evaluating ambient artificial intelligence documentation: effects on work efficiency, documentation burden, and patient-centered care. J. Am. Med. Inform. Assoc. 2025:ocaf180.

9 Shah SJ, Crowell T, Jeong Y, et al. Physician perspectives on ambient ai scribes. *JAMA Netw*. Open 2025;8:e251904.

10 Shah KP, Johnson KB. The ambient ai scribe revolution—early gains and open questions. *JAMA Netw*. Open 2025;8:e2534982.

11 Pearlman K, Wan W, Shah S, et al. Use of an ai scribe and electronic health record efficiency. *JAMA Netw*. Open 2025;8:e2537000.

12 Rotenstein LS, Holmgren AJ, Thombley R, et al. Changes in clinician time expenditure and visit quantity with adoption of artificial intelligence–powered scribes: a multisite study. JAMA 2026;335:1408–17.

13 Husa RA, Haggerty J, Nute AW, et al. Ambient artificial intelligence use and clinician documentation burden, productivity, and efficiency. *JAMA Netw*. Open 2026;9:e2615762.

14 Holmgren AJ, Fenton CL, Thombley R, et al. Ambient artificial intelligence scribes and physician financial productivity. *JAMA Netw*. Open 2026;9:e2553233.

15 Chandrasekaran R, Moustakas E. Patient attitudes toward ambient artificial intelligence scribes in clinical care: insights from a cross-sectional study. J. Am. Med. Inform. Assoc. 2026;33:263–72.

16 Feng J, Phillips RV, Malenica I, et al. Clinical artificial intelligence quality improvement: towards continual monitoring and updating of ai algorithms in healthcare. Npj Digit. Med. 2022;5:66.

17 Thai K, Emi B, Masrour E, et al. EditLens: quantifying the extent of ai editing in text. 2025.

18 Guo Y, Hu D, Yang Z, et al. Clinicians’ rationale for editing ambient ai–drafted clinical notes: persistent challenges and implications for improvement. J. Am. Med. Inform. Assoc. 2026:ocag059.

19 Anderson TN, Mohan V, Dorr DA, et al. Evaluating the quality and safety of ambient digital scribe platforms using simulated ambulatory encounters. Mayo Clin. Proc. Digit. Health 2025;3:100292.

20 Shaw SD, Nave G. Thinking—fast, slow, and artificial: how ai is reshaping human reasoning and the rise of cognitive surrender. 2026.

21 Abdelwanis M, Alarafati HK, Tammam MMS, et al. Exploring the risks of automation bias in healthcare artificial intelligence applications: a bowtie analysis. J. Saf. Sci. Resil. 2024;5:460–69.

22 Guo Y, Hu D, Yang Z, et al. What do clinicians edit in ambient ai-drafted clinical documentation? a qualitative content analysis. J. Am. Med. Inform. Assoc. 2026:ocag073.

23 Moramarco F, Papadopoulos Korfiatis A, Perera M, et al. Human evaluation and correlation with automatic metrics in consultation note generation. In: Muresan S, Nakov P, Villavicencio A, eds. Proceedings of the 60th Annual Meeting of the Association for Computational Linguistics (Volume 1: Long Papers) Dublin, Ireland: Association for Computational Linguistics 2022 :5739–54.

24 Yujian L, Bo L. A normalized levenshtein distance metric. IEEE Trans. Pattern Anal. Mach. Intell. 2007;29:1091–95.

25 Zhang T, Kishore V, Wu F, et al. BERTScore: evaluating text generation with bert. 2020.

26 Chang E, Sung S. Use of snomed ct in large language models: scoping review. JMIR Med. Inform. 2024;12:e62924.

27 Qiu P. Introduction to statistical process control. CRC press; 2013.

28 Feng J, Xia F, Singh K, et al. Not all clinical ai monitoring systems are created equal: review and recommendations. NEJM AI 2025;2:AIra2400657.

