## Supplement for "A Framework to Monitor Editing of Artificial Intelligence-Generated Medical Documentation"

**SUPPLEMENTARY MATERIAL 1: Validation of clinical edit intensity as a measure of clinically meaningful edits**

**Annotator Task Instructions**

For this annotation task, a number of HTML documents and an excel file will be shared with you. Each HTML document represents a section (either HPI or A&P) of an encounter note. These sections were initially generated by the AI scribe Abridge and subsequently edited by the clinician. The information shared with you for each section includes the section text and an encounter ID.

You will see text in three formats:

- ~~AI-generated text that was removed by the clinician appears like this~~
- AI-generated text that was not edited appears like this
- Text that was manually added by the clinician appears like this

There are therefore two types of edits, ~~removals~~ and additions. For this task, you only need to evaluate these changes.

**Your task is to determine if these edits introduced changes that could be considered clinically meaningful. Possible answers are ‘Yes’, ‘No’, or ‘Unsure’.**

Edits can be considered clinically meaningful when one or more of the edits lead to a change in clinical meaning. Examples of clinically meaningful edits are:

-> She presents with worsened symptoms ~~probably due to her IBD~~

-> When she injured her ~~right~~ left knee

-> I think that this is lumbar radiculopathy

Rephrasing or reformatting, as well as removing irrelevant or negated content should not be considered clinically meaningful. Examples of not clinically meaningful edits are:

-> The patient ~~denies chest pain~~ reports no chest pain

-> Lapse in care following his previous PCP’s retirement (Dr. Stan Smith)

-> ~~He is accompanied by his mother today~~

-> ~~No mention of prior imaging~~

Some edits may be ambiguous, but they should not be automatically considered clinically meaningful. Examples of ambiguous edits are:

-> Changes in dates should not be considered meaningful, unless they are very relevant to	the visit

-> Adding or removing medications or numeric lab results, unless change information	significantly. Bear in mind that removing a lab or a medication should be considered	meaningful. For example:

Reports borderline high blood pressure (135/85) -> Not meaningful

She takes ~~advil~~ ibuprofen as needed -> Meaningful

-> Any changes that are not relevant to the ongoing visit

Complete the excel file by filling out the column ‘clinically meaningful’ associated with each individual encounter ID. Please answer ‘Yes’, ‘No’, or ‘Unsure’ exclusively. Please use the ‘Unsure’ label sparingly.

You may also add observations in the ‘comments’ column, especially for encounters labeled ‘Unsure’.

**Validation of clinical edit intensity as a measure of clinically meaningful edits**

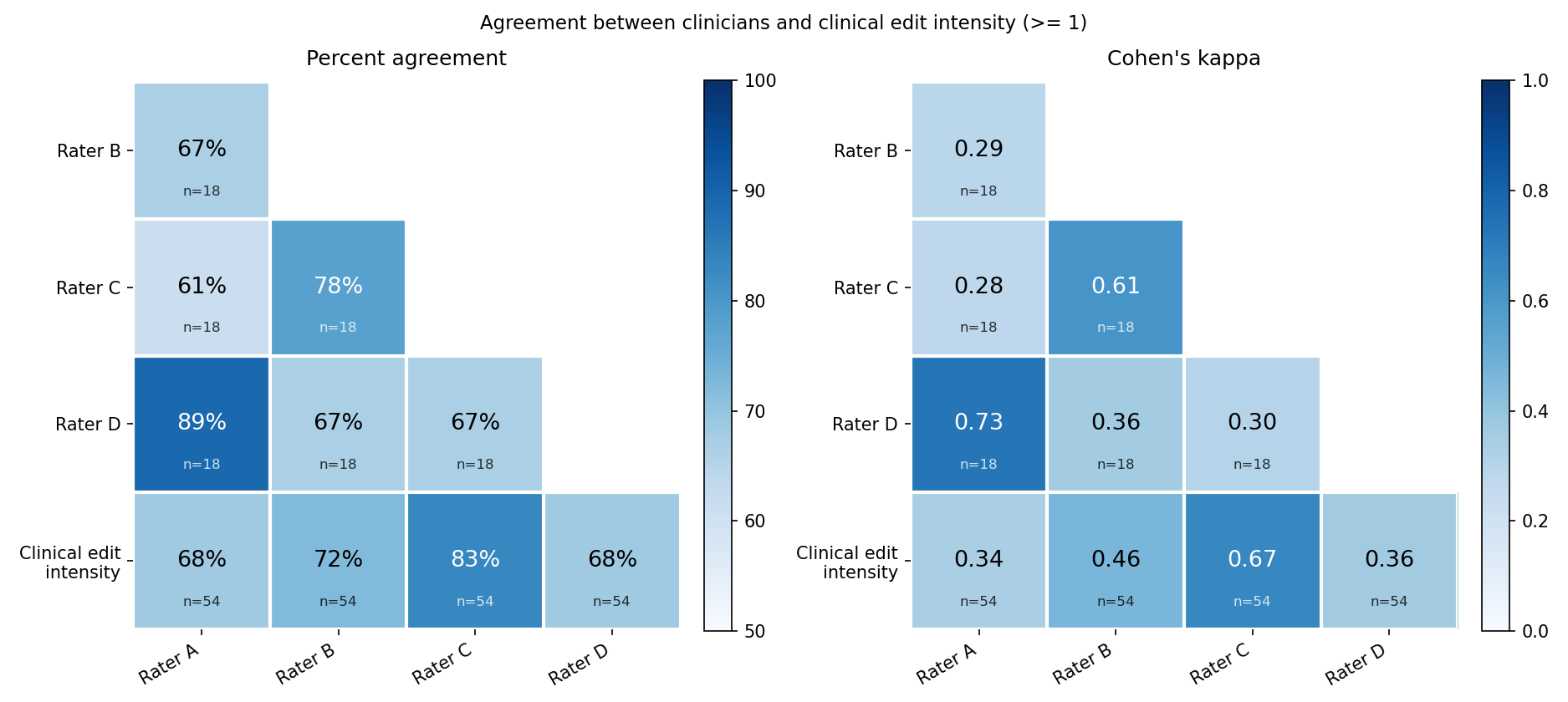

**Figure S1.** Percent agreement (left) and Cohen’s κ (right) between all annotator pairs and annotator-model pairs.

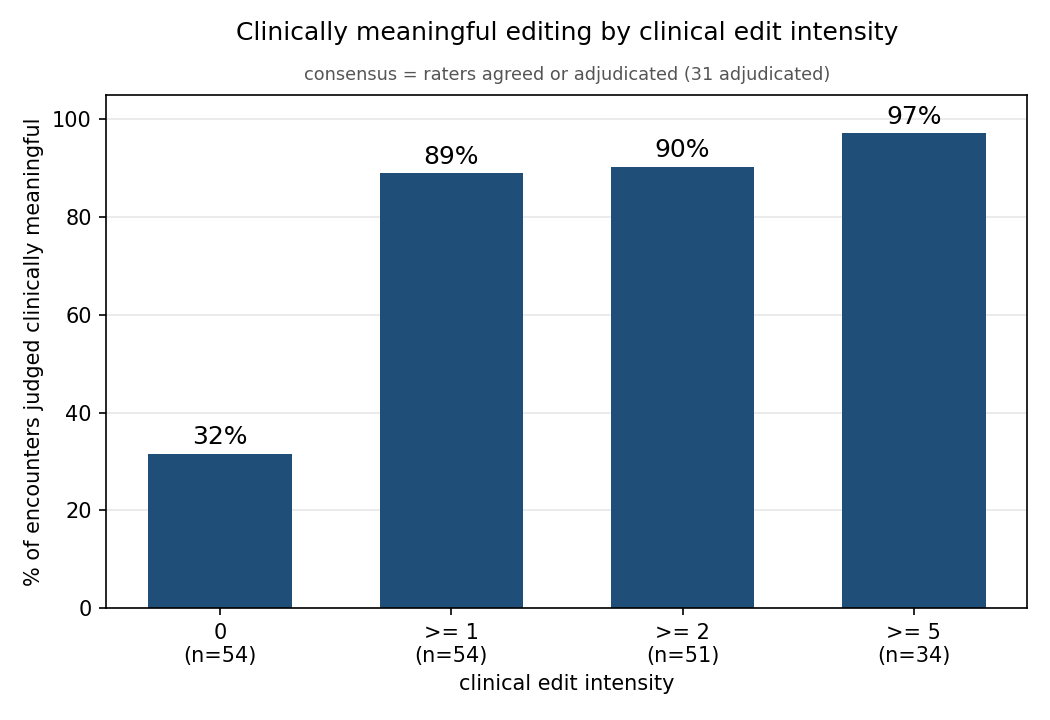

**Figure S2.** Positive predictive value of clinical edit intensity as a measure of clinically meaningful editing at different thresholds when compared to adjudicated annotations.

| **Table S1**. Breakdown of positive predictive value (PPV), negative predictive value (NPV), percent agreement and Cohen’s κ between clinical edit intensity and adjudicated annotations | | | | |
| --- | --- | --- | --- | --- |
| Section | PPV (Clinical edit intensity >=1) | NPV (Clinical edit intensity 0) | Percent Agreement | Cohen’s κ |
| HPI | 85.2 | 63.0 | 74.1 | 0.48 |
| A&P | 92.6 | 74.1 | 83.3 | 0.67 |
| Overall | 88.9 | 68.5 | 78.7 | 0.57 |

**SUPPLEMENTARY MATERIAL 2: Feasibility analysis of using the framework to monitor system-level changes in editing**

**EWMA Control Charts for New Patient Encounters and Encounters requiring an Interpreter**

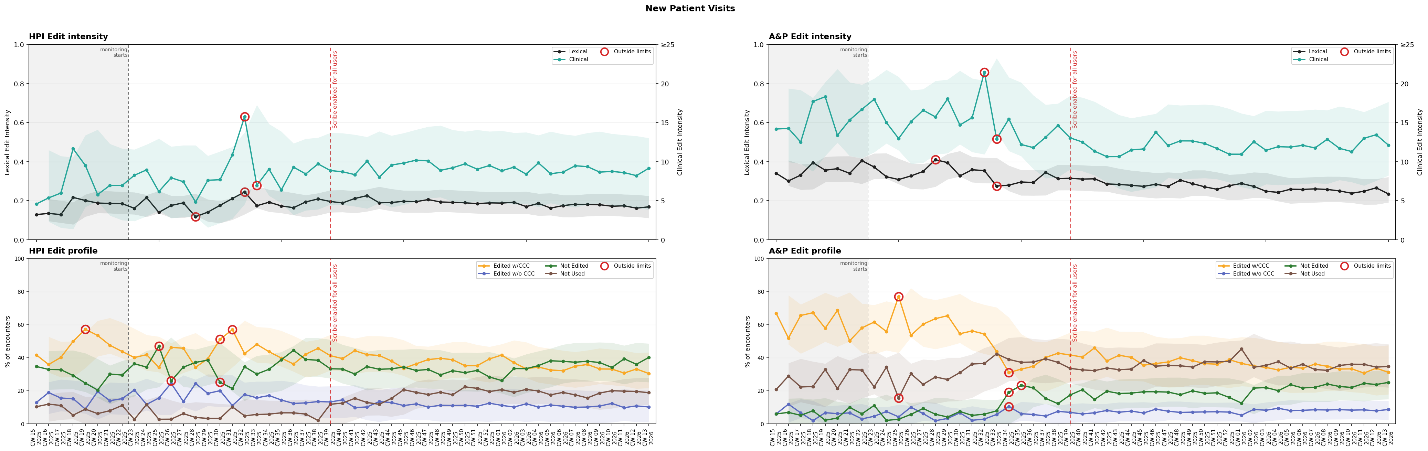

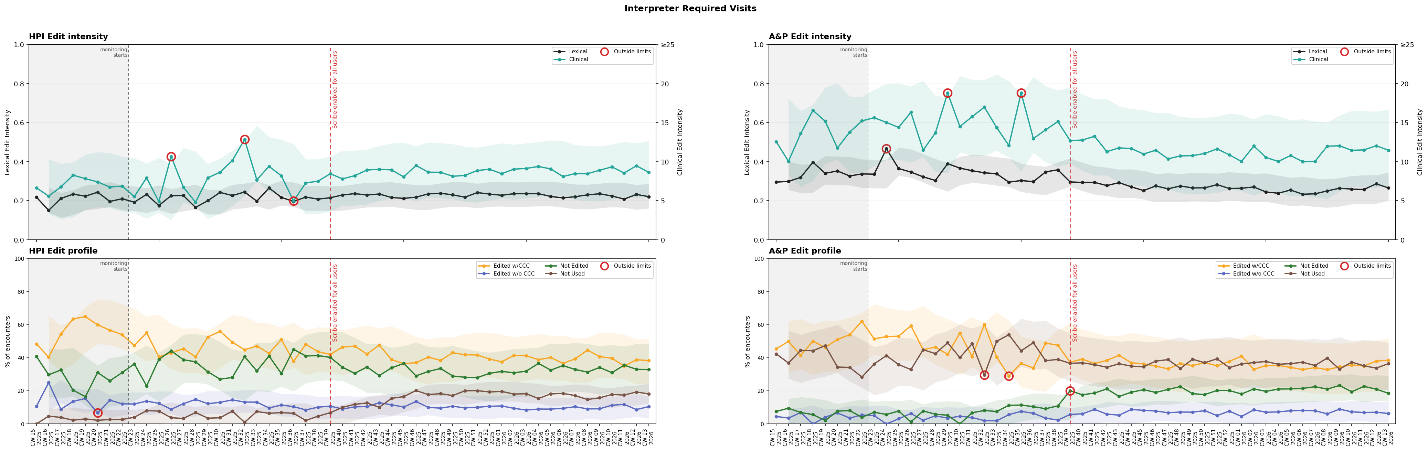

**Figure S3.** Exponentially weighted moving average (EWMA) run charts of editing dimensions and profiles for new patient encounters (top) and encounters requiring an interpreter (bottom). The unit of measure in the x axis is calendar week (CW). ‘Not Used’ includes all encounters in the ‘Not used/Alternative Workflow’ profile. Run chart parameters: $\lambda=0.75, \rho=2.3$. CCC: Clinical Concept Change.

**EWMA Control Charts with Different Values of Lambda and Rho**

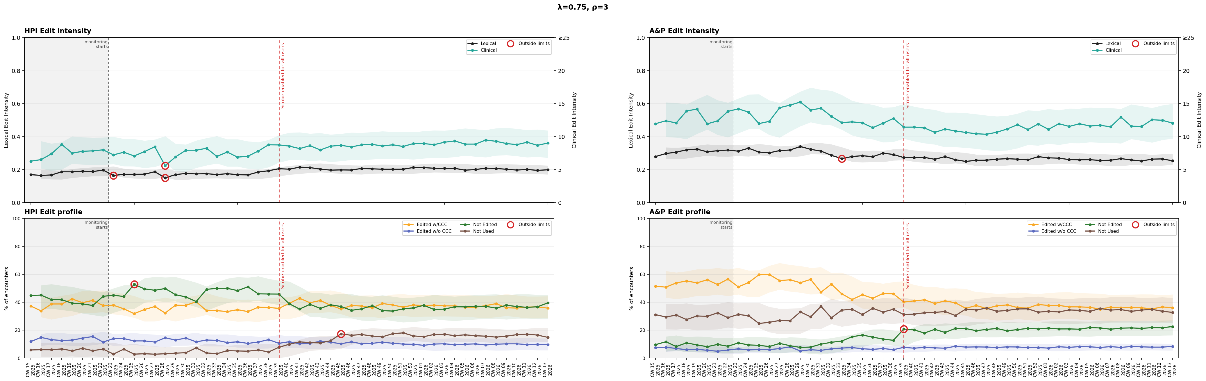

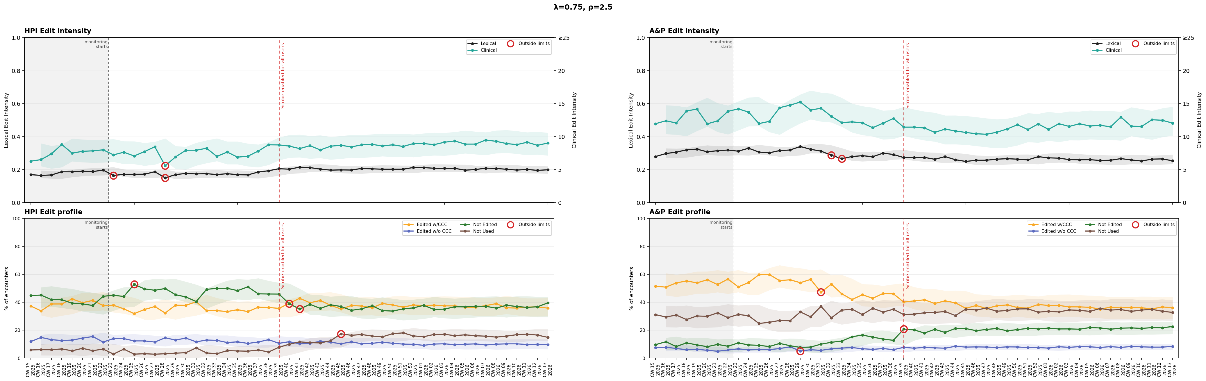

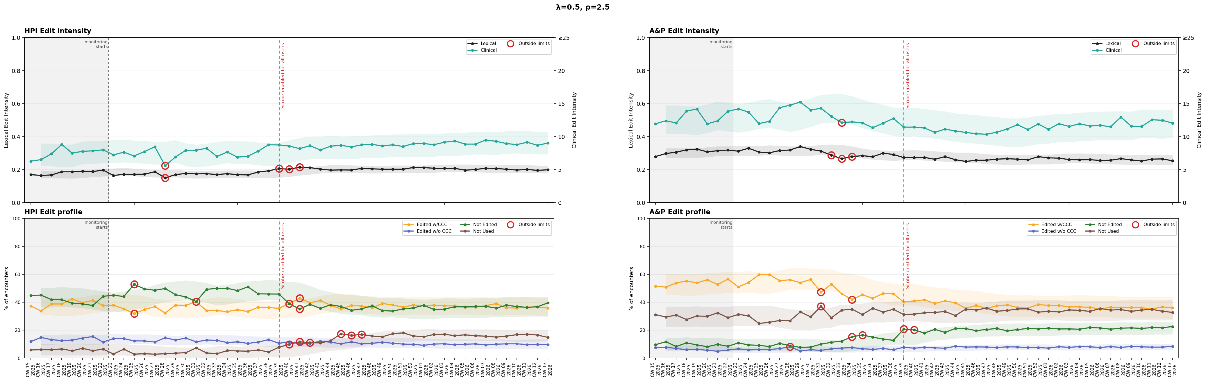

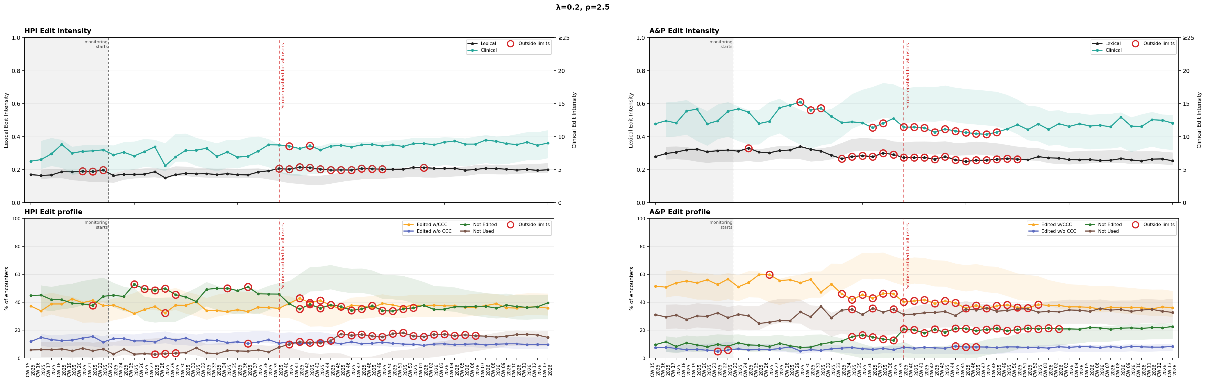

**Figure S4.** Exponentially weighted moving average (EWMA) run charts of editing dimensions and profiles with different values of $\lambda$ and $\rho$. ‘Not Used’ includes all encounters in the ‘Not used/Alternative Workflow’ profile. CCC: Clinical Concept Change. The unit of measure in the x axis is calendar week (CW).

**Relative Changes Needed to Exceed EWMA Control Limits for Different Values of Lambda and Rho**

| **Table S2**. Relative change (%) needed to exceed exponentially weighted moving average control chart limits for different values of lambda and rho for each measure and section. | | | | | | |
| --- | --- | --- | --- | --- | --- | --- |
| Lambda | Rho | Section | Measure | Last Value | Relative Change Needed, Lower (%) | Relative Change Needed, Upper (%) |
| 0.75 | 2.3 | HPI | Lexical | 0.199 | -10.8 | 10 |
| 0.75 | 2.3 | HPI | Clinical | 9.02 | -18.7 | 16.4 |
| 0.75 | 2.3 | HPI | Edited w/o CCC | 9.649 | -32.4 | 35.3 |
| 0.75 | 2.3 | HPI | Edited w/CCC | 35.855 | -15.6 | 17.7 |
| 0.75 | 2.3 | HPI | Not Edited | 39.594 | -20.5 | 13.3 |
| 0.75 | 2.3 | HPI | Not Used | 14.901 | -19.5 | 31.5 |
| 0.75 | 2.3 | A&P | Lexical | 0.253 | -8.4 | 12.3 |
| 0.75 | 2.3 | A&P | Clinical | 12.038 | -15.3 | 18.1 |
| 0.75 | 2.3 | A&P | Edited w/o CCC | 8.456 | -25 | 18.7 |
| 0.75 | 2.3 | A&P | Edited w/CCC | 36.082 | -19.9 | 20.5 |
| 0.75 | 2.3 | A&P | Not Edited | 22.669 | -20.2 | 16.1 |
| 0.75 | 2.3 | A&P | Not Used | 32.792 | -19.3 | 23.1 |
| 0.2 | 2.5 | HPI | Lexical | 0.199 | -8.4 | 20.2 |
| 0.2 | 2.5 | HPI | Clinical | 9.02 | -25.6 | 21.4 |
| 0.2 | 2.5 | HPI | Edited w/o CCC | 9.649 | -32.4 | 55.1 |
| 0.2 | 2.5 | HPI | Edited w/CCC | 35.855 | -13.6 | 30.1 |
| 0.2 | 2.5 | HPI | Not Edited | 39.594 | -33.7 | 10.2 |
| 0.2 | 2.5 | HPI | Not Used | 14.901 | -32.1 | 39.9 |
| 0.2 | 2.5 | A&P | Lexical | 0.253 | -8.6 | 21.7 |
| 0.2 | 2.5 | A&P | Clinical | 12.038 | -31.4 | 12.2 |
| 0.2 | 2.5 | A&P | Edited w/o CCC | 8.456 | -41.8 | 18 |
| 0.2 | 2.5 | A&P | Edited w/CCC | 36.082 | -19.1 | 32.6 |
| 0.2 | 2.5 | A&P | Not Edited | 22.669 | -40.3 | 8.8 |
| 0.2 | 2.5 | A&P | Not Used | 32.792 | -19.8 | 32.8 |
| 0.5 | 2.5 | HPI | Lexical | 0.199 | -10.9 | 11.9 |
| 0.5 | 2.5 | HPI | Clinical | 9.02 | -19.8 | 18.1 |
| 0.5 | 2.5 | HPI | Edited w/o CCC | 9.649 | -32 | 39 |
| 0.5 | 2.5 | HPI | Edited w/CCC | 35.855 | -15.4 | 20 |
| 0.5 | 2.5 | HPI | Not Edited | 39.594 | -24.7 | 10.7 |
| 0.5 | 2.5 | HPI | Not Used | 14.901 | -16.6 | 38.2 |
| 0.5 | 2.5 | A&P | Lexical | 0.253 | -8.7 | 14.6 |
| 0.5 | 2.5 | A&P | Clinical | 12.038 | -17 | 18.8 |
| 0.5 | 2.5 | A&P | Edited w/o CCC | 8.456 | -28.9 | 18 |
| 0.5 | 2.5 | A&P | Edited w/CCC | 36.082 | -21.2 | 21.2 |
| 0.5 | 2.5 | A&P | Not Edited | 22.669 | -24.1 | 14.8 |
| 0.5 | 2.5 | A&P | Not Used | 32.792 | -17.3 | 26.5 |
| 0.75 | 2.5 | HPI | Lexical | 0.199 | -11.8 | 10.9 |
| 0.75 | 2.5 | HPI | Clinical | 9.02 | -20.3 | 18 |
| 0.75 | 2.5 | HPI | Edited w/o CCC | 9.649 | -35.3 | 38.3 |
| 0.75 | 2.5 | HPI | Edited w/CCC | 35.855 | -17 | 19.2 |
| 0.75 | 2.5 | HPI | Not Edited | 39.594 | -21.9 | 14.7 |
| 0.75 | 2.5 | HPI | Not Used | 14.901 | -21.7 | 33.7 |
| 0.75 | 2.5 | A&P | Lexical | 0.253 | -9.3 | 13.2 |
| 0.75 | 2.5 | A&P | Clinical | 12.038 | -16.7 | 19.6 |
| 0.75 | 2.5 | A&P | Edited w/o CCC | 8.456 | -26.9 | 20.6 |
| 0.75 | 2.5 | A&P | Edited w/CCC | 36.082 | -21.7 | 22.3 |
| 0.75 | 2.5 | A&P | Not Edited | 22.669 | -21.7 | 17.7 |
| 0.75 | 2.5 | A&P | Not Used | 32.792 | -21.2 | 24.9 |
| 0.75 | 3 | HPI | Lexical | 0.199 | -14 | 13.2 |
| 0.75 | 3 | HPI | Clinical | 9.02 | -24.1 | 21.8 |
| 0.75 | 3 | HPI | Edited w/o CCC | 9.649 | -42.7 | 45.6 |
| 0.75 | 3 | HPI | Edited w/CCC | 35.855 | -20.7 | 22.8 |
| 0.75 | 3 | HPI | Not Edited | 39.594 | -25.6 | 18.4 |
| 0.75 | 3 | HPI | Not Used | 14.901 | -27.2 | 39.3 |
| 0.75 | 3 | A&P | Lexical | 0.253 | -11.6 | 15.5 |
| 0.75 | 3 | A&P | Clinical | 12.038 | -20.3 | 23.2 |
| 0.75 | 3 | A&P | Edited w/o CCC | 8.456 | -31.7 | 25.4 |
| 0.75 | 3 | A&P | Edited w/CCC | 36.082 | -26.1 | 26.6 |
| 0.75 | 3 | A&P | Not Edited | 22.669 | -25.7 | 21.7 |
| 0.75 | 3 | A&P | Not Used | 32.792 | -25.8 | 29.6 |
